# Infantile Colic in Karachi: Exploring Maternal Awareness, Attitude and Management Approaches

**DOI:** 10.1101/2024.07.09.24310028

**Authors:** Syed Rohan Ali, Moosa Abdur Raqib, Kiran Mehtab, Munir Nafees, Malik Hamdan Tafheem

## Abstract

Infantile colic, characterized by severe abdominal pain and excessive crying, significantly impacts both infants and their parents. This study examined maternal awareness, knowledge, and attitudes towards colic, with a focus on socio-demographic factors. Conducted over eight months with a sample of 400 infants in Karachi, the research employed structured questionnaires and clinical evaluations. Statistical analysis revealed significant variations in colic occurrence based on age (p < 0.001), maternal age (p = 0.005), and educational background (p = 0.001). Gender differences in digestive issues (p = 0.02) and responses to crying based on gestational age were also notable. Additionally, educational status significantly affected perceptions of colic severity (p = 0.000) and its impact on parental mental health (p = 0.03). These findings highlight the necessity for tailored healthcare strategies that consider familial contexts and educational interventions to enhance understanding and management of infantile colic. Future research should explore the influence of gut microbiota and probiotics, aiming to develop empathetic and evidence-based approaches to improve outcomes for affected families.

**Objective:** This study aimed to provide updated insights into the epidemiology of infantile colic in Karachi, contributing to targeted healthcare interventions and policies aimed at alleviating colic-related distress among infants and their families in urban settings

## Introduction

Infantile colic, marked by acute abdominal pain in infants, is mainly associated with gastrointestinal issues, though its exact cause is still unknown. The symptoms, including excessive crying and fussiness, cause significant distress for both infants and their parents. This study aims to evaluate mothers’ awareness, knowledge, and attitudes towards infantile colic to better understand its causes. By examining socio-demographic factors such as maternal age and gender, the study seeks to identify potential differences in awareness. Understanding these maternal perspectives is essential, as they directly affect the child’s well-being and overall family dynamics. This research aims to contribute to a thorough understanding and develop targeted interventions for managing infantile colic in Pakistan.

According to the recent definition by Hyman, colicky infants cry consistently during the evening at around the same time each day for at least one week, yet they remain otherwise healthy.

Fortunately, infantile colic is typically short-lived: it usually starts at about two weeks of age and improves by the fourth month.^1^ Various management strategies, including behavioral, dietary, pharmacological, and alternative interventions, are discussed. However, due to a lack of large, high-quality randomized controlled trials, none of these therapies are strongly recommended.^2^

The study also summarizes the behavioral and somatic consequences of infantile colic into childhood.

Although infantile colic usually does not lead to long-term problems, the persistent crying can cause parental frustration, postpartum depression, frequent doctor visits, and even child abuse. Colic undoubtedly induces stress within the family and diminishes the quality of life for both the family and the infant. Some evidence suggests that colicky babies may be at increased risk for recurrent abdominal pain in childhood or more frequent functional gastrointestinal disorders (FGIDs) and mood disorders in adulthood compared to healthy infants without colic. It has also been suggested that infants with colic have a higher risk of developing migraines. However, well-designed long-term, prospective observational studies are needed to confirm the few reports on potential links between infantile colic and future health outcomes.

## Methods

A prospective cohort cross-sectional study was conducted on a sample of 400 infants, selected through non-probability purposive sampling from the National Institute of Child Health, Dr. Ruth K. M. Pfau, Civil Hospital Karachi, and Sindh Employees Social Security Institution (SESSI) Hospital Landhi Karachi. The study took place over an eight-month period from October 2023 to May 2024. Informed verbal consent was obtained from the parents, and a structured questionnaire was distributed among them. A pilot study was conducted to assess the validity of the questionnaire. To ensure representation from various socioeconomic backgrounds, a diverse sample of infants aged 0-12 months was enrolled from multiple healthcare facilities across Karachi using a stratified random sampling approach. The validated structured questionnaire collected comprehensive data on colic symptoms, maternal demographics, and household characteristics. Clinical examinations by healthcare providers further validated colic diagnoses and excluded other potential causes of infant distress. Statistical analysis was performed using SPSS Version 22, with a 95% confidence interval, a 5% margin of error, and a significance level set at 0.05. Ethical considerations included obtaining informed consent from participants and ensuring the confidentiality of collected data. This study aimed to provide updated insights into the epidemiology of infantile colic in Karachi, contributing to targeted healthcare interventions and policies aimed at alleviating colic-related distress among infants and their families in urban settings.

## Result

The study examined infant abdominal colic across various factors, revealing significant findings. Differences in colic occurrence were significant across age ranges (p < 0.001), with variability noted in reported frequencies for different age groups. Maternal age (p = 0.005) and educational background (p = 0.001) also significantly influenced colic reports. Family size showed significant variations in reported colic frequencies (p = 0.011), while timing of colic varied significantly across age groups (p = 0.004). Maternal education significantly affected perceptions of colic causes (p = 0.034) and severity as a health issue (p = 0.000).

Gender differences were significant in reported digestive issues (p = 0.02) and occurrence of a high-pitched sound (p = 0.008) among infants. Responses to infant crying varied significantly by gestational age for home remedies (p = 0.001), OTC medications (p = 0.001), and seeking medical advice (p = 0.004). Family size influenced opinions on awareness about colic (p = 0.001). Birth weight showed significant associations with perceptions of colic severity (p = 0.035).

Educational status significantly impacted perceptions of colic as a health issue (p = 0.000) and its impact on parental mental health (p = 0.03). Overall, these findings underscore the multifaceted influences on how colic is reported, perceived, and managed across different demographic factors.

## Discussion

Infantile colic is a common yet challenging condition affecting infants worldwide, characterized by excessive crying and fussiness. Understanding its prevalence and management requires consideration of various demographic factors and parental perceptions, as highlighted by recent data analysis. Significant variations were observed in the reported occurrence of infant abdominal colic across different age ranges, with statistical significance noted in distributions among infants aged 0-6 months (p = 0.000). This suggests that the manifestation of colic symptoms varies significantly during early infancy, potentially influenced by developmental stages and physiological changes in newborns.

Moreover, the frequency of colic pain reported by mothers varied notably based on the number of children in families (p = 0.011). Families with fewer children tended to report higher frequencies of daily colic pain, reflecting potential differences in caregiving practices and family dynamics that may impact infant health outcomes. These findings underscore the need for tailored healthcare strategies that consider familial contexts and support mechanisms to alleviate the burden of colic on parents and caregivers.

Educational status emerged as a critical determinant of maternal perceptions regarding infantile colic. Mothers with higher educational attainment were more likely to perceive colic pain as a serious health issue compared to those with lower educational backgrounds (p = 0.000). This disparity highlights the influence of maternal education on health literacy and the interpretation of infant health symptoms, indicating a need for targeted educational interventions to enhance understanding and management practices among all caregivers.

Furthermore, the association between birth weight and perceptions of colic pain severity was statistically significant (p = 0.035). Parents of lighter infants tended to perceive colic as a more serious health concern compared to those with heavier infants, reflecting potential biases in parental perceptions related to infant health outcomes. Healthcare providers should address these biases through comprehensive education and support initiatives to ensure equitable care for all infants affected by colic, regardless of birth weight or perceived severity of symptoms.

In addition to demographic influences, the study also examined parental responses based on gestational birth age, revealing significant differences in treatment-seeking behaviors (p-values: home remedy use = 0.001, over-the-counter medication = 0.001, doctor’s advice = 0.004). Term infants were more likely to receive home remedies and over-the-counter medications compared to pre-term and post-term infants, suggesting variations in healthcare utilization based on gestational age and perceived vulnerability to colic-related discomforts.

The impact of infantile colic on parental mental health also varied significantly according to educational status (p = 0.03). Parents with lower educational attainment reported a greater perceived impact of colic on their mental well-being compared to those with higher education levels, underscoring the need for targeted support services and mental health interventions for vulnerable caregivers. Addressing these disparities requires holistic approaches that integrate psychological support and coping strategies tailored to the unique needs of affected families.

In conclusion, the findings from this study provide valuable insights into the multifaceted nature of infantile colic, emphasizing the influence of demographic factors, parental perceptions, and healthcare-seeking behaviors on its prevalence and management. These insights underscore the importance of tailored interventions that consider familial contexts, educational backgrounds, and birth weight categories to optimize care and support for infants and their families. Future research should continue to explore these dynamics to enhance our understanding of colic etiology, treatment efficacy, and the long-term implications for child development and family well-being. By addressing these factors comprehensively, healthcare providers can improve outcomes and quality of life for infants and families affected by this common yet impactful condition.

## Conclusion

Based on our analysis, effective management of infantile colic should inegrate evidence-based strategies alongside empathetic support. Clinicians are advised to adopt a multifaceted approach, combining non-pharmacological interventions with parental education to address this common yet distressing condition. Non-pharmacological methods such as the “five S’s” offer a safe and economical means to comfort colicky infants, prioritizing parental involvement and emotional reassurance without the risks associated with pharmacological treatments. It is crucial to emphasize the self-limiting nature of colic, typically resolving by four to six months of age, thereby alleviating parental anxiety through informed reassurance.

Further research is imperative to deepen our understanding and enhance treatment outcomes for infantile colic. Ongoing investigation into probiotics and their mechanisms, as well as the influence of gut microbiota on colic, represents promising avenues for future study. Tailored interventions targeting diverse infant populations should be explored to account for variability in symptoms and responsiveness to treatment. Rigorous placebo-controlled trials with larger sample sizes are essential to bolster the current evidence base, ensuring that clinical practices remain grounded in robust scientific findings and ultimately improving the management of infantile colic globally.

## Data Availability

will provided access on request

## Declarations

## Author Contributions

- **Concept & Design of Study:** Syed Rohan Ali
- **Drafting:** Moosa Abdur Raqib
- **Data Analysis:** Syed Rohan Ali
- **Revisiting Critically:** Syed Rohan Ali
- **Final Approval of Version:** Moosa abdur raqib and Kiran Mehtab

SYED ROHAN ALI, MOOSA ABDUR RAQIB, KIRAN MEHTAB, MUNIR NAFEES, MALIK HAMDAN TAFHEEM played crucial roles in refining the content, conducting a final round of editing, and presenting the article in a structured and balanced format, ensuring it was ready for publication.

## IRB Reference No

IRB-65/04/ LCMD/08/2022

## Ethics Approval

Ethics approval was granted by the IRB Committee LCMD, Karachi.

## Consent

Verbal consent was obtained from the participants.

## Data Availability

Data was collected by the investigator. If required, data and materials access can be granted.

---

**TABLES.**
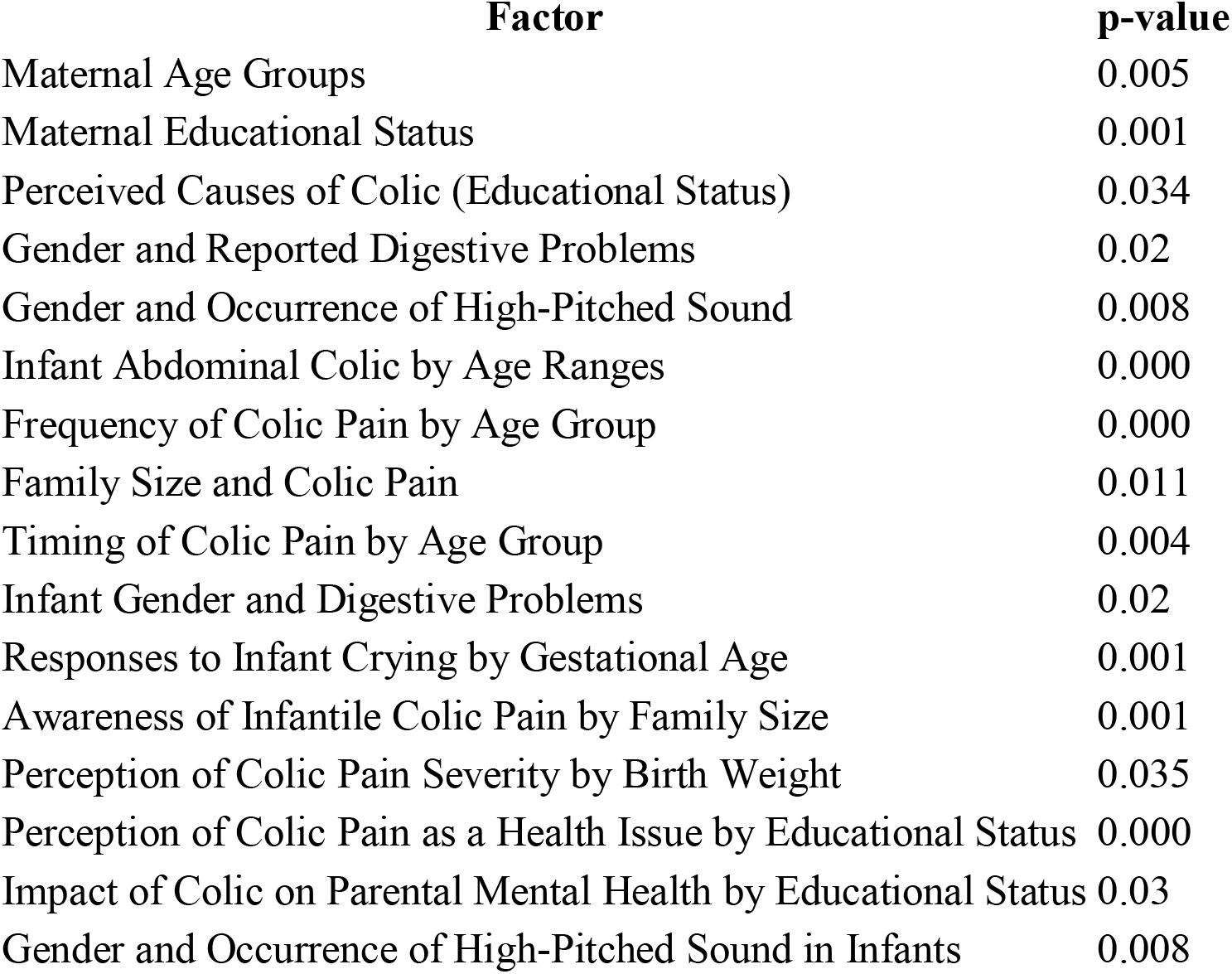
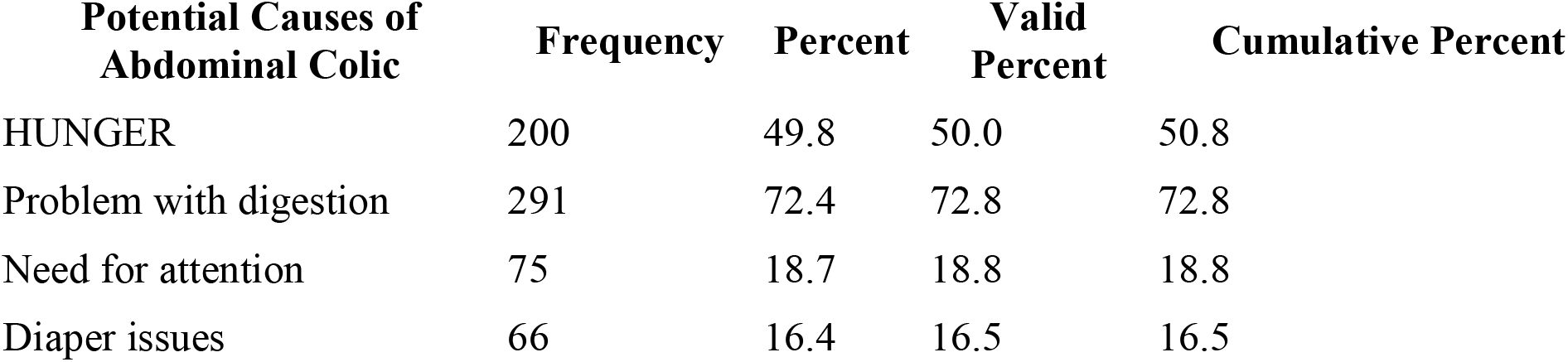
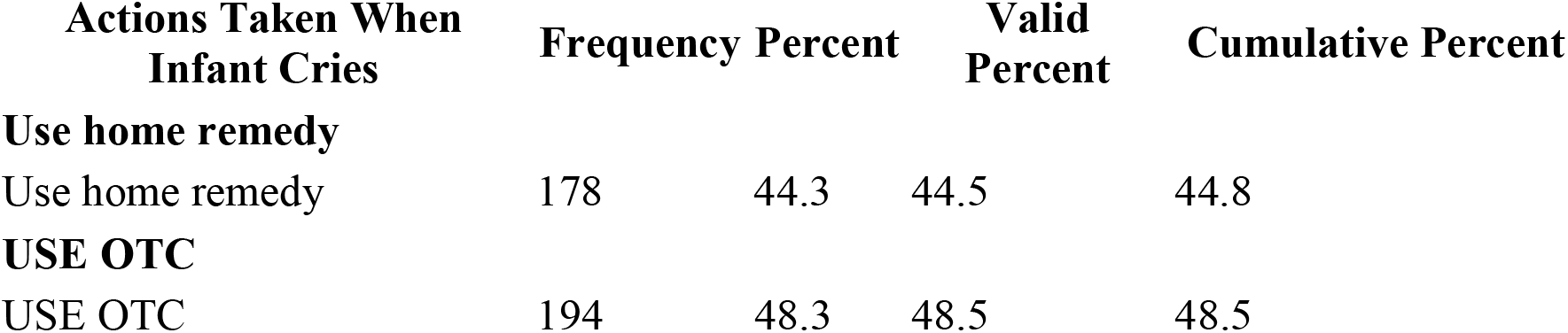
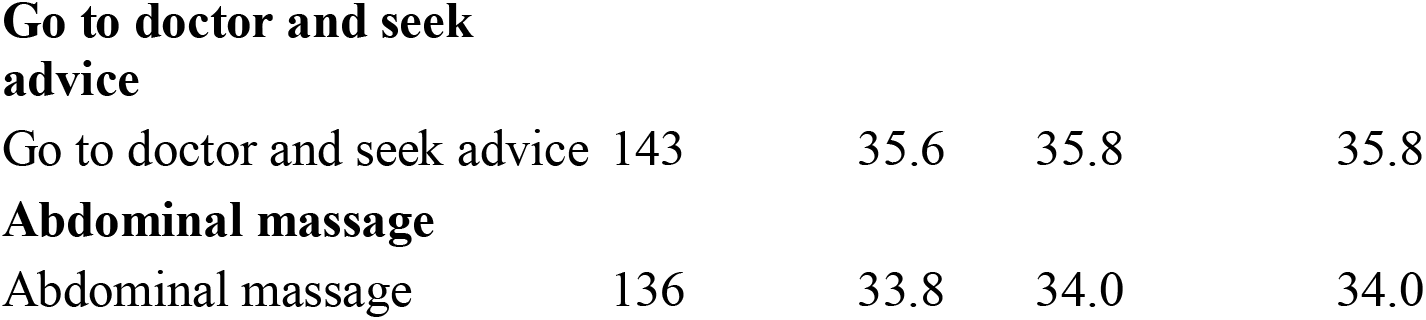
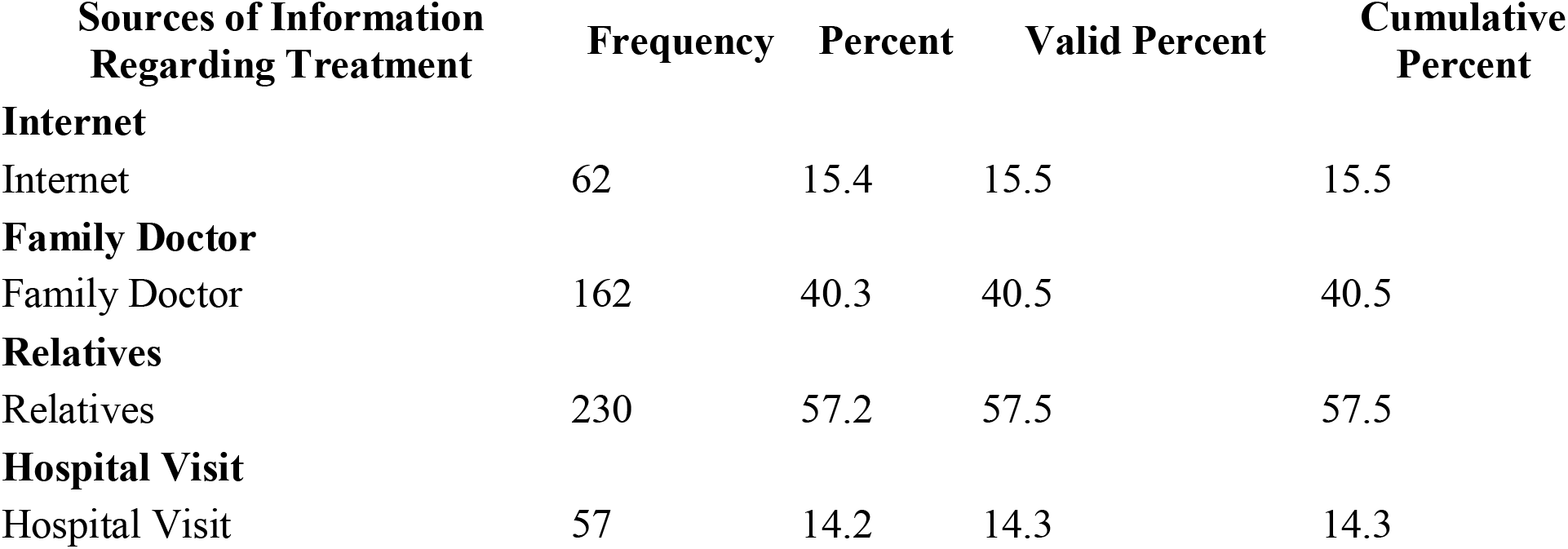
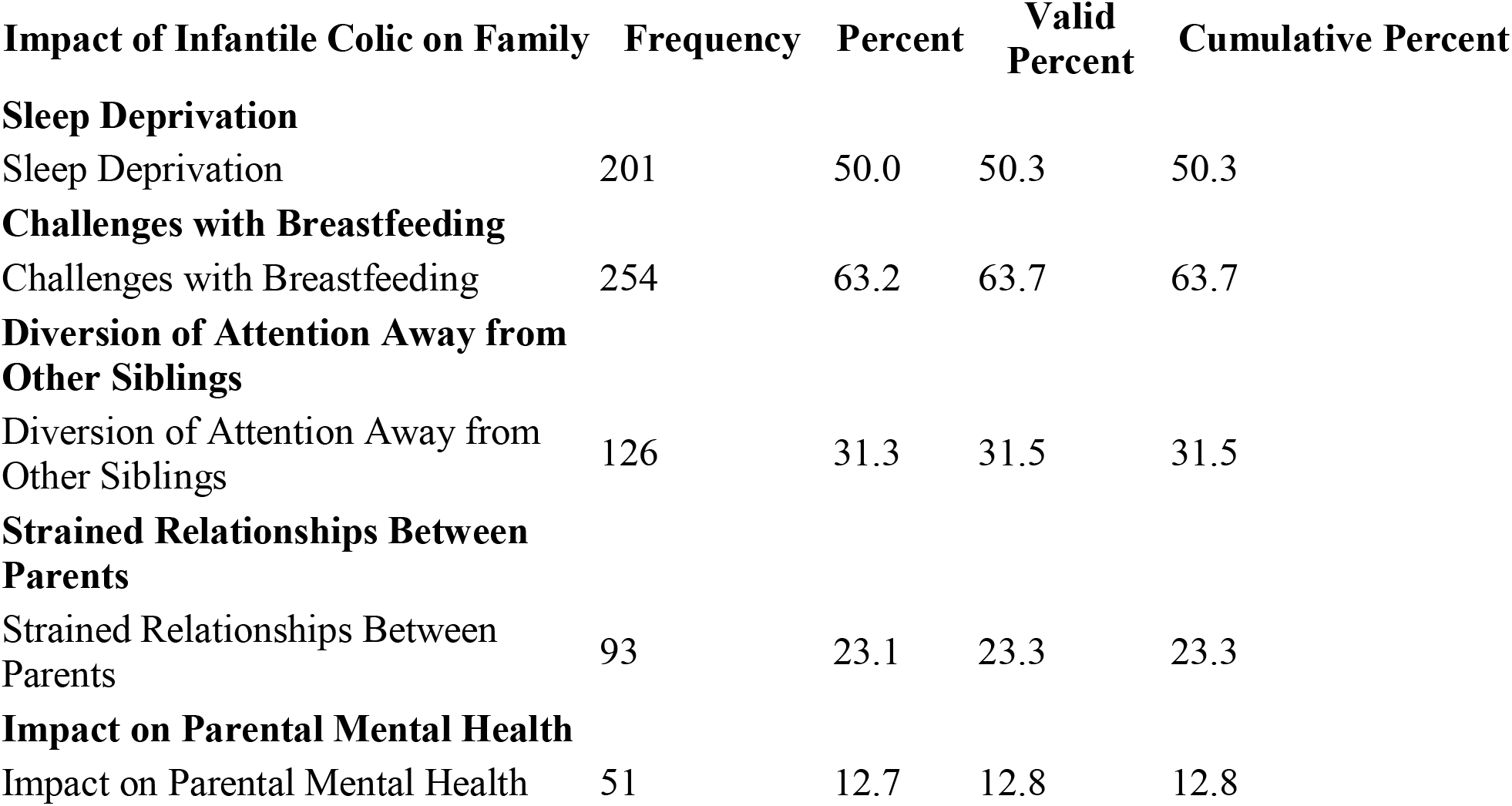
Summary of p-Values for Various Factors Related to Infantile Colic.

